# Early evidence for the safety of certain COVID-19 vaccines using empirical Bayesian modeling from VAERS

**DOI:** 10.1101/2021.06.10.21258589

**Authors:** Chris von Csefalvay

**Affiliations:** Starschema Inc., Arlington, VA

## Abstract

The advent of vaccines against SARS-CoV-2 ushered in an unprecedented global response to COVID-19, with the largest and most ambitious mass vaccination campaign in human history. The scale of this effort means that safety signals suggesting adverse effects may only be detectable using passive reporting. This paper examines reports to the CDC/FDA’s VAERS system in the first six months of 2021, using an empirical Bayesian model with a gamma Poisson shrinker to identify potential safety signals from COVID-19 vaccines currently on the U.S. market. Based on this preliminary data, it is concluded that the COVID-19 vaccine’s safety significantly exceeds that of previously marketed vaccines, and other than a known risk of thrombotic events, no safety signals of concern emerge.

## 1 Introduction

The introduction of vaccines against SARS-CoV-2 has lent the response to the COVID-19 a new string to its bow. In the world’s largest mass vaccination campaign to date,[1–3] over two million doses of SARS-CoV-2 vaccines have been administered by the end of May 2021 – over 300 million of these just in the United States. As with any medical intervention, it is indispensible for public confidence that any potential risks be identified and corrected early. Adverse events following immunisation (AEFIs) have been documented in the context of every known vaccine, and are mostly benign (such as pyrexia, transient nonspecific malaise and injection site discomfort). In the context of the COVID-19 vaccines, identifying particular clinically significant safety signals is made more difficult by the scale of the pandemic. The logistics of active surveillance within a pandemic are daunting at best, limiting us primarily to deriving insights from passive surveillance.

In passive surveillance of AEFIs, persons who experience AEFIs volunteer information about their experience. In the United States, the leading system for this purpose is VAERS, jointly maintained by the CDC and the FDA. Passive surveillance, however, suffers from both over-and underreporting: while some patients do not report their AEFIs (especially if these are mild and transient), many who report various ailments do so regardless of a determination of causality. VAERS was intentionally designed to be ‘over-inclusive’: anyone can submit reports, and there is no requirement for evidence to claim a particular AEFI. Moreover, the structure of VAERS allows for a wide range of information to be included as ‘symptoms’, including tests carried out and tests with normal results. There is also an inherent awareness bias: patients with more severe AEFIs are more likely to make an effort to report their symptoms than those experiencing mild reactions, skewing the relative reporting rate to over-estimate the real occurrence of serious AEFIs.[4]

The COVID-19 global vaccination campaign adds another layer of complexity onto a field already fraught with difficulties. Because of the sheer number of vaccines administered, even an exceedingly rare side effect may create a large enough number of affected patients. In order to address the growing concern of vaccine hesitancy,[5–7] it is crucial to analyse and present AEFIs in the context of the overall number of vaccinations. Estimating risks using appropriate metrics and comparing it to can play a very significant role in dispelling misinformation and supporting evidence-based decision-making by patients and physicians alike.[8,9]

This paper focuses on the early evidence from the first six months of COVID-19 vaccination in the United States, from 1 January to 28 May 2021, and concludes that the safety record of the COVID-19 vaccines appears to be solid at this time based on the available evidence from VAERS. Using a Bayesian framework for isolating potential safety signals, data submitted to VAERS is compared to other vaccines during the same time period, concluding with an evaluation of the COVID-19 vaccines’ overall safety.

## 2 Methods

### 2.1 Data set

Data for this study was obtained from VAERS on 06 June, 2021. At the time of retrieval, the data set included reports received on or before 28 May, 2021. Data was retrieved using the CDC bulk download site.

### 2.2 Processing

Data was processed using R 4.1.0 [10]. Upon import, data was destructured from VAERS’s multi-event schema, where multiple putative AEFIs are included in a single line, to a single-event schema using `reshape2`.[11]

### 2.3 Metrics

One of the most widely used metrics to identify possible safety signals is the Proportional Reporting Ratio (PRR).[12] For the *m* × *n* matrix *D* of *m* adverse events and *n* drugs, where *D*_*i,j*_ (*i* ∈*m, j* ∈*n*), the PRR of side effect *i* in the presence of the drug *j* is defined as

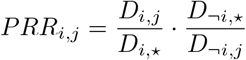

The PRR commends itself by relative mathematical simplicity and ease of implementation, but is subject to a disproportional reporting bias. In other words, the PRR does not indicate whether a certain side effect is more or less frequent compared to another, or with another drug. In particular, it does not reflect relative risk. It often eludes even trained professionals that the correct interpretation of *PRR*_*i,j*_ is not the relative probability that a certain adverse effect will be reported with this particular drug compared with the reference drugs. Thus, a *PRR*_*anaphylaxis,j*_ of 3.0 does not indicate that anaphylaxis is three times more likely with *j* than any other drug. Instead, it indicates that the probability of reporting anaphylaxis rather than any other event with *j* is three times higher than the probability of reporting anaphylaxis rather than any other event with other drugs.[13]

A better indicator of possible safety signals is the empirical Bayesian geometric mean (EBGM) or modified DuMouchel’s method.[14] Since its first publication in 1999, this method has been widely used in analysing ‘market basket’ type problems – that is, identifying combinations of elements on each axis that occur with unusual frequency, where a Bayesian baseline is calculated through an expectation prior.[15–17]

The EBGM approach builds on the relative reporting ratio *R*_*rep*_ (occasionally also *RR*), defined as 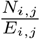, where *Ni, j* is the actual number of reported instances of the adverse effect *i* given the drug *j*. One would thus expect a value of 1.0 if no association existed, i.e. if rows and columns were independent from each other. Higher values would thus increasingly militate away from the null hypothesis and towards an association between *i* and *j*.

One of the deficiencies of the *R*_*rep*_ metric is that for low-expectancy low-occurrence issues, a single integer occurrence (which may well be entirely accidental) may, in the face of a small real valued expectancy value, result in a misleadingly high *R*_*rep*_ (e.g. *E*_*i,j*_ = 0.05, *N*_*i,j*_ = 1 yields an *R*_*rep*_ of 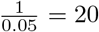.) DuMouchel’s work expands on this by using a Poisson likelihood for actual counts, in which *N*_*i,j*_ = *Poisson*(*µ*_*i,j*_). [14] This affords us the ability to calculate the metric

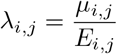

for a prior on *λ*_*i,j*_ being drawn from a mixture of two gamma distributions. The posterior distribution of *λ*_*i,j*_, specifically, is the mixture of two gamma distributions parametrised by the shape and scale variables {*α*_1_, *β*_1_} and {*α*_2_, *β*_2_} . Consequently, the two distributions are parametrised by

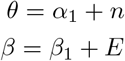

and

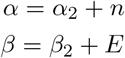

with the parameter 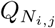 being the mixture fraction (i.e. the likelihood that *λ*_*i,j*_ was drawn from the first gamma distribution of the posterior). Consequently, the posterior of *λ* is a probabilistic-Bayesian representation of *R*_*rep*_ (and thus amenable to similar canons of interpretation), but with more stable results for low-expectancy low-occurrence events.

### 2.4 Computation

Computation was carried out using the openEBGM[18] package under R 4.1.0.[10] Data was stratified by gender (male, female and unknown) and age group. Age groups were aggregated into four bins: <25, 25-44, 45-64 and over 65 years of age. The Cartesian product of the two stratum variables yielded 15 strata.

For the estimation of hyperparameter vector *θ* = (*α*_1_, *β*_1_, *α*_2_, *β*_2_, *Q*), the nonlinear Newton minimisation function `stats::nlm` was used, with initialisation weights of *α*_1_ = 0.2, *β*_1_ = 0.1, *α*_2_ = 2.0, *β*_2_ = 4.0 and *Q* = 0.333.

The computation was carried out in two separate runs. First, the data was examined over vaccine types (VAERS variable `VAX_TYPE`), e.g. `FLU3` for all trivalent influenza vaccines and `COVID19` for all COVID-19 vaccines. Then, the same methodology, including fitting separate values for 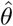, was applied to the data over individual vaccines (VAERS variable `VAX_NAME`). In both cases, the same stratification was used.

In addition to the EBGM values, the mixture fraction *Q*_*n*_ of the posterior probability distribution was estimated using the formula described by Eqn. 6 in DuMouchel (1999).[14] Finally, the `quantBisect` function of the `openEBGM` package was used to estimate 5th and 95th percentiles, thereby providing a two-sided 10% confidence margin.

## 3 Results

### 3.1 Absolute results

The absolute results of the analysis shows COVID-19 vaccines as a group have a remarkably favourable safety profile. Of the 12,477 vaccine-symptom combinations for COVID-19 vaccines, only 24 had an EBGM mean value exceeding 2.0, commonly regarded as the lower bound for identifying a safety signal. Of the 24, 7 (29.17%) are entries for tests conducted and/or normal results and 3 (12.5%) are productor administration-inherent reports (e.g. temperature excursion during product storage). Besides the generic entry for ‘adverse drug reaction’, the only identifiable clinical pictures recorded with an EBGM value exceeding 2.0 were deep vein thrombosis (DVT), gaze palsy, thrombosis and central venous sinus thrombosis.

The mean EBGM for COVID vaccines was 0.9936 (*σ* = 0.1629), indicating a highly favourable safety profile. When analysed as a group, there were very few side effects even mildly above an EBGM of 1.00, indicating that the vaccine behaved as predicted. The proportionally highest proportional reported rate was a papular rash (*PRR* = 8.55), while the largest absolute numbers of reports were non-specific symptoms that are common AEFIs and indicate immune activation. These include headaches (60,490 reports), pyrexia (49,459 reports) and chills (47,650 reports). Of 1,242,557 distinct reports of symptoms from COVID-19 vaccines during the examined period, only 3,769 involved death. It is important to note at this juncture that these reports are not verified, nor is causal attribution performed. The number of deaths (regardless of reporting confidence and lack of attribution) must be seen in the context of the fact that these reports arose from over 300 million doses of vaccination, putting the reporting likelihood at approx. one report of a death for every 79,500 doses administered.

### 3.2 Versus other vaccine types

Compared to other vaccine categories, COVID-19 vaccines have the lowest mean EBGM, at almost exactly 1.00. There is a risk that the overall much higher number of vaccines administered, and as such the higher number of reports (during the period under examination, 1,252,858 distinct symptom reports were made, with 1,242,557, or 99.18%, of these being for a COVID-19 vaccine) presents some distortion, enhancing the central tendency of data on the COVID-19 vaccine. Nonetheless, Table 1 provides a convincing comparison that attests to the safety of the COVID-19 vaccines vis-a-vis other vaccine types.

**Table 1:**
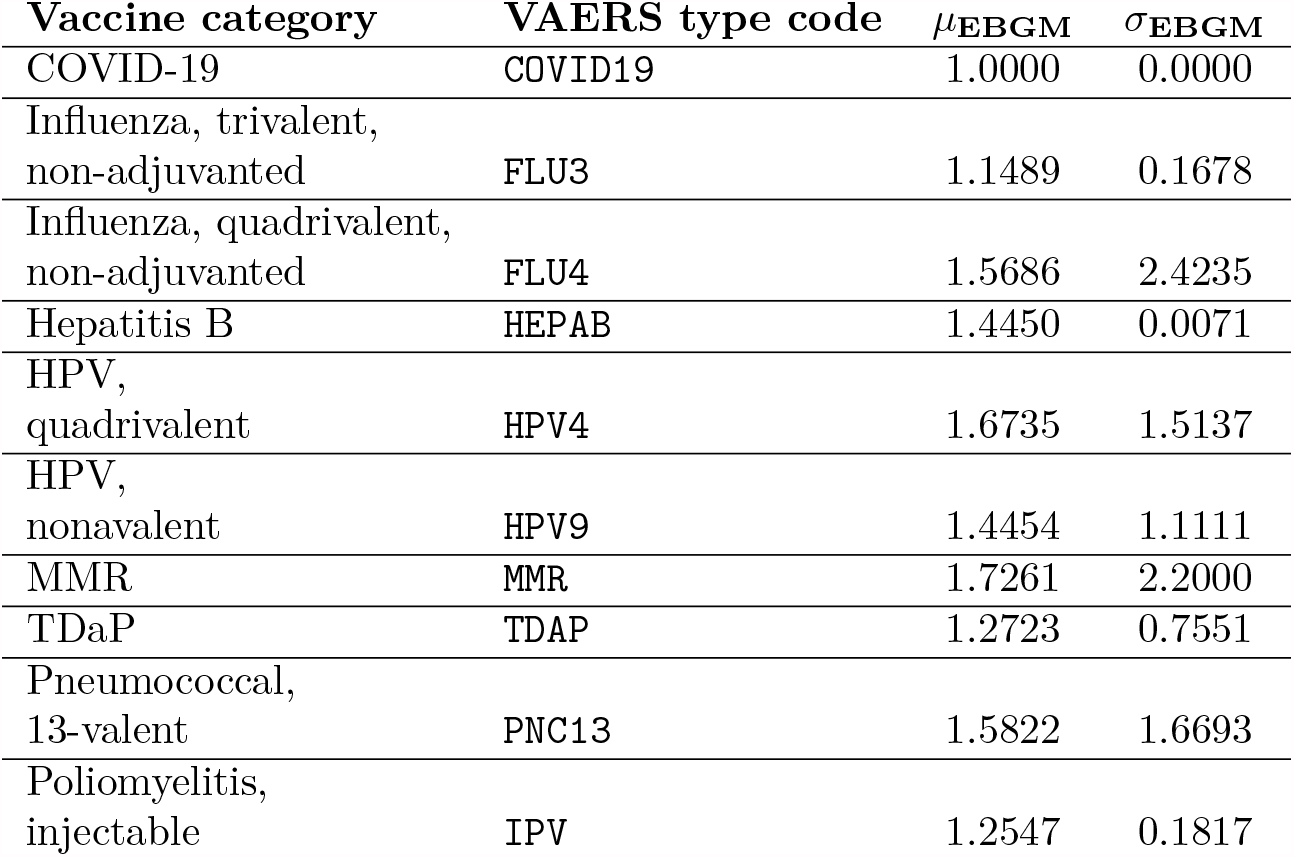
Mean EBGM (*µ*_*EBGM*_) and standard deviation (*s*_*EBGM*_) of the COVID-19 vaccine and a selection of vaccines in common clinical use within the United States

## 4 Discussion

As both Table 1 and Figure 1 indicate, the VAERS data between 01 January and 28 May 2021 shows a favourable side effect profile for the COVID-19 vaccine when compared to other commonly used vaccines.

**Figure 1:**
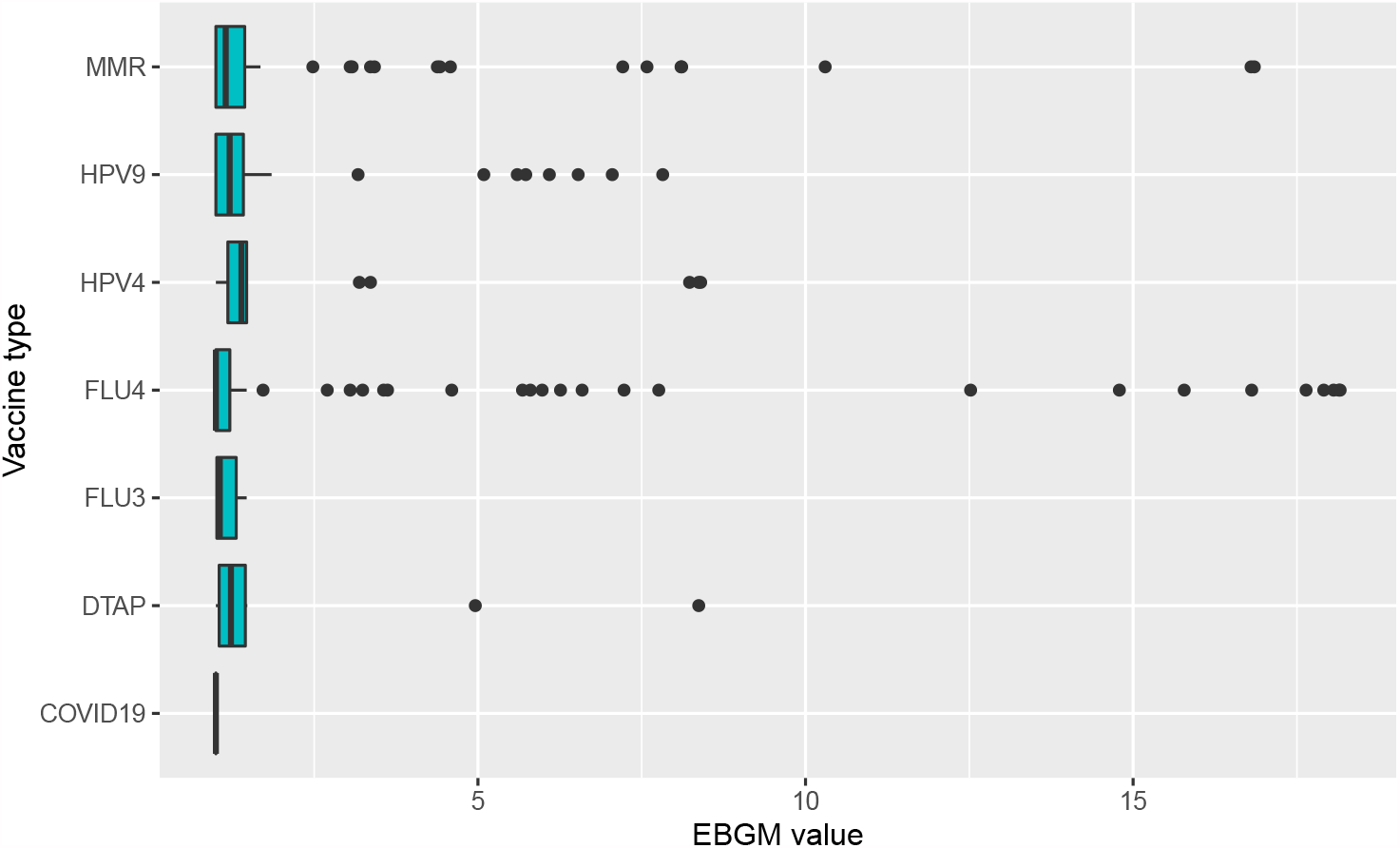
Comparison of EBGM values for the COVID-19 vaccine and a selection of vaccines in common clinical use within the United States.

From an analytical perspective, the disproportionate volume of COVID-19 vaccines when compared with all other vaccines poses a challenge. Typically, vaccines are administered to an age limited spectrum of the population – for instance, children typically receive their second TDaP vaccine between the ages of 4 and 6. Emergency vaccination campaigns that encompass the entire population may pose unique analytical problems. Thus, for instance, the number of reported instances of a certain AEFI may give a misleading indication of that AEFI’s prevalence. The Bayesian approach underlying the EBGM algorithm, which uses informative priors and focuses on the relative difference in the posterior, controls for these discrepancies effectively.

Based on reports between 01 January and 28 May 2021, the short to midterm safety of COVID-19 vaccines appear to be firmly established. AEFIs appear primarily to be the typical concomitants of immune activation (arthralgia, pyrexia and non-specific malaise), with a potentially clinically significant but very rare indication of thrombotic events. The clinical experience so far appears to confirm this.[19,20] Long-term surveillance efforts will require more data, as it will certainly accrue over time, and may call for these findings be confirmed in view of new information. For the time being, however, the safety of COVID-19 vaccines appears settled.

## Data Availability

All data sources are publicly available from the CDC. The source code is available under https://github.com/chrisvoncsefalvay/vaers-covid.

## References

[1] Sanjeet Bagcchi. The world’s largest COVID-19 vaccination campaign. The Lancet Infectious Diseases, 21(3):323, 2021.

[2] Yingzhu Li, Rumiana Tenchov, Jeffrey Smoot, Cynthia Liu, Steven Watkins, and Qiongqiong Zhou. A comprehensive review of the global efforts on COVID-19 vaccine development. ACS Central Science, 7(4): 512–533, 2021.

[3] Edouard Mathieu, Hannah Ritchie, Esteban Ortiz-Ospina, Max Roser, Joe Hasell, Cameron Appel, Charlie Giattino, and Lucas Rodés-Guirao. A global database of COVID-19 vaccinations. Nature Human Behaviour, pages 1–7, 2021.

[4] Frederick Varricchio, John Iskander, Frank Destefano, Robert Ball, Robert Pless, M Miles Braun, and Robert T Chen. Understanding vaccine safety information from the vaccine adverse event reporting system. The Pediatric infectious disease journal, 23(4):287–294, 2004.

[5] Jagdish Khubchandani, Sushil Sharma, James H Price, Michael J Wiblishauser, Manoj Sharma, and Fern J Webb. COVID-19 vaccination hesitancy in the United States: a rapid national assessment. Journal of Community Health, 46(2):270–277, 2021.

[6] Patrick Peretti-Watel, Valérie Seror, Sébastien Cortaredona, Odile Launay, Jocelyn Raude, Pierrea Verger, Lisa Fressard, François Beck, Stéphane Legleye, Olivier l’Haridon, et al. A future vaccination campaign against COVID-19 at risk of vaccine hesitancy and politicisation. The Lancet Infectious Diseases, 20(7):769–770, 2020.

[7] Mohammad S Razai, Umar AR Chaudhry, Katja Doerholt, Linda Bauld, and Azeem Majeed. Covid-19 vaccination hesitancy. bmj, 373, 2021.

[8] Sahil Loomba, Alexandre de Figueiredo, Simon J Piatek, Kristen de Graaf, and Heidi J Larson. Measuring the impact of COVID-19 vaccine misinformation on vaccination intent in the uk and usa. Nature Human Behaviour, 5(3):337–348, 2021.

[9] Jon Roozenbeek, Claudia R Schneider, Sarah Dryhurst, John Kerr, Alexandra LJ Freeman, Gabriel Recchia, Anne Marthe Van Der Bles, and Sander Van Der Linden. Susceptibility to misinformation about COVID-19 around the world. Royal Society open science, 7(10):201199, 2020.

[10] R Core Team. R: A Language and Environment for Statistical Computing. R Foundation for Statistical Computing, Vienna, Austria, 2021.

[11] Hadley Wickham. reshape2: Flexibly reshape data: a reboot of the reshape package. R package version, 1(2), 2012.

[12] Stephen JW Evans, Patrick C Waller, and S Davis. Use of proportional reporting ratios (PRRs) for signal generation from spontaneous adverse drug reaction reports. Pharmacoepidemiology and drug safety, 10(6):483– 486, 2001.

[13] Nicholas Moore, Gillian Hall, Miriam Sturkenboom, Ron Mann, Rajaa Lagnaoui, and Bernard Begaud. Biases affecting the proportional reporting ratio (prr) in spontaneous reports pharmacovigilance databases: the example of sertindole. Pharmacoepidemiology and drug safety, 12(4):271–281, 2003.

[14] William DuMouchel. Bayesian data mining in large frequency tables, with an application to the fda spontaneous reporting system. The American Statistician, 53(3):177–190, 1999.

[15] June S Almenoff, William DuMouchel, L Allen Kindman, Xionghu Yang, and David Fram. Disproportionality analysis using empirical Bayes data mining: a tool for the evaluation of drug interactions in the post-marketing setting. Pharmacoepidemiology and Drug Safety, 12(6):517–521, 2003.

[16] Rave Harpaz, William DuMouchel, Paea LePendu, and Nigam H Shah. Empirical bayes model to combine signals of adverse drug reactions. In Proceedings of the 19th ACM SIGKDD international conference on Knowledge discovery and data mining, pages 1339–1347, 2013.

[17] Hyesung Lee, Ju Hwan Kim, Young June Choe, and Ju-Young Shin. Safety surveillance of pneumococcal vaccine using three algorithms: Disproportionality methods, empirical bayes geometric mean, and tree-based scan statistic. Vaccines, 8(2):242, 2020.

[18] Travis Canida and John Ihrie. openEBGM: An R implementation of the Gamma-Poisson Shrinker data mining model. R J., 9(2):499, 2017.

[19] Tom T Shimabukuro. COVID-19 vaccine safety update. 2021.

[20] Kerry J Welsh, Jane Baumblatt, Wambui Chege, Ravi Goud, and Narayan Nair. Thrombocytopenia including immune thrombocytopenia after receipt of mRNA COVID-19 vaccines reported to the Vaccine Adverse Event Reporting System (VAERS). Vaccine, 39(25):3329–3332, 2021.

